# Knowledge, attitude, and practices of Community pharmacists about COVID-19: A cross-sectional survey in two provinces of Pakistan

**DOI:** 10.1101/2020.05.22.20108290

**Authors:** Khayal Muhammad, Muhammad Saqlain, Ataullah Hamdard, Muhammad Naveed, Muhammad Farooq Umer, Siraj Khan, Muhammad Kamran, Haroon Rashid, Sohail Kamran, Muhammad Ijaz Khan, Faiz Ullah Khan, Yaseen Hussain, Zakir Khan

**Author notes:** Corresponding author: Zakir Khan. Current affiliation: Institute of Health Sciences, Department of Pharmacology (Pharmacovigilance), Cukurova University, Adana, Turkey. Previous affiliation: Department of Pharmacy, Quaid-i-Azam University, Islamabad, Pakistan. ORCID: http://orcid.org/0000-0003-1365-548X.

## Abstract

Amidst to current Coronavirus infectious disease 2019 (COVID-19) pandemic, the international pharmaceutical federation stated that pharmacists being a part of the healthcare system had a crucial role in the management cycle of COVID-19 outbreak. The purpose of this study was to assess the knowledge, attitude and practice of community pharmacists, to snapshot their current preparedness and awareness regarding COVID-19. An online survey was conducted among a sample of 393 community pharmacists from two provinces; Punjab and Khyber-Pakhtunkhwa, Pakistan during a period of strict lockdown (10^th^ to 30^th^ April 2020). A validated (Cronbach alpha= 0.077) self-administered questionnaire comprised of five sections (Demographics, source of information, knowledge, attitude, and practice) was used for data collection. Logistic regression was applied to find potential factors associated with good knowledge, attitude, and practice by using SPSS version 21. Of total 393 participants, 71.5% (n=281) had good knowledge, 44% (n=175) had positive attitude and 57.3% (n=225) had good practice regarding COVID-19. Social media (45.29%, n=178) was reported as the main source to seek information regarding COVID-19. Results revealed that the age of ≥26 years, Ph.D. degree level, and good knowledge were the substantial determinants (P<0.05) of a good attitude. Similarly, community pharmacist who had an experience of >5 years, hold a Ph.D. degree, good knowledge and good attitude had higher odds of good practice compared to reference categories (P<0.05). The findings demonstrated that the majority of community pharmacists had good knowledge, but had a poor attitude and practice towards the COVID-19. This study also highlighted the disparity in some aspects of knowledge, attitude, and practice that must be addressed in future educational, awareness, and counselling programs.

## 1. Introduction

China became the center of an infectious outbreak called Coronavirus disease 2019 (COVID-19) in December 2019, which gained intense attention not only in China but also globally. Cases of COVID-19 were no longer limited to China, with the increasing number of cases and widening geographical spread raising grave concerns about the future trajectory of the outbreak [1]. COVID-19 virus is a zoonotic beta-coronavirus similar to the viruses that are responsible to cause severe acute respiratory syndrome (SARS) and the middle east respiratory syndrome (MERS). COVID-19 can spread via close person-to-person contact, coughing and sneezing with a symptom of fever, dry cough, and shortness of breath and may sometimes cause severe diseases, such as pneumonia, respiratory distress, and death in some cases [1, 2].

The SARS epidemic, which started in 2002, ended up numbering about 8,000 infected people and 770 dead, contributing to global costs of up to $100 billion. In comparison, MERS was characterized by fewer infections but stronger nosocomial outbreaks and a slightly higher mortality rate (approximately 35 %) and has not been eliminated to date with more than 90,000 confirmed cases and more than 2,500 deaths [3]. The world health organization (WHO) declared the COVID-19 as an international pandemic public health emergency [4]. The WHO data on the 18th of May 2020 showed more than 4618800 confirmed and 311847 death cases due to the COVID-19 throughout the globe [5].

The health care system of different countries started effective planning to cope with the COVID-19 pandemic. Pharmacists are an important part of the health care system, and their role is critical in completing the management cycle of coronavirus outbreak [2]. Pharmacists being a member of the health care team can play an important role in disease management and outbreak surveillance [6]. Globally, in the current situation of lockdown, pharmacies are among the few shops or places that are kept open for public services and therefore community pharmacists are the first point of contact to fulfill the community’s healthcare needs. Community pharmacists are vital health care providers and remained on the frontline during the COVID-19 outbreak for public health by serving as direct points of access for their patients and playing an important role during the outbreak in infection control as well as patient counseling, care, and support [7].

The international pharmaceutical federation (FIP) emphasized on the effective role of pharmacists in the community for preventing the spread of COVID-19 [8]. Community pharmacists often act as a reliable information source for individuals having concerns or needing information and advice regarding ailments. Moreover, pharmacists are readily available at community pharmacies and accessible to the general population [7, 8]. Community pharmacists across the world have a central role to play an effective role in COVID-19 preventive measures and action. There is an urgent need to support their critical role by health care policymakers of every country for better patient care [6].

In the near future, the confirmed and fatality cases of COVID-19 is expected to increase significantly in limited resources due to the weak health care system and facilities [9]. Pakistan is a limited resource country with the most vulnerable geographical location for this pandemic as it shares borders with the most affected countries such as China and Iran. According to the WHO report on May 18, 2020, Pakistan has 42125 confirmed cases and 903 deaths due to the COVID-19 [5]. Pakistani health care system faced a lack of basic health facilities, recommended policies, unavailability of proper medical equipment, financial crisis, and a limited number of health care professionals to handle such outbreak [10]. There is limited availability of community pharmacists in Pakistan and also their role is not properly defined [11]. The fight against COVID-19 is continuing in Pakistan and involvement of community pharmacist’s effective role is a need of time to fight with this global threat.

Experience gained from previous SARS outbreak in 2002 suggests that knowledge and attitude towards contagious infections are linked with a level of panic affection amid the community, which can further complicate chances to inhibit the transmission of the infection [12]. Knowledge, attitude, and practice survey provide a format to evaluate existing programs and to identify effective strategies for behavior change in society. Currently, there is scarce information regarding the awareness level of community pharmacists in Pakistan. Knowledge, attitude, and practice survey offer a convenient plan to check the existing programs and to find out adequate approaches for behavior change in the community. Presently, there is limited information regarding the awareness level of the Pakistani community pharmacists. To promote COVID-19 outbreak management in Pakistan, it is very important to find out the awareness of community pharmacists about COVID-19 in current serious situation. In the present research, we determined the knowledge, attitude and practice (KAP) of community pharmacists towards COVID-19 during the fast rise period of the pandemic outbreak.

## 2. Methods

### 2.1. Study design

A cross-section online survey was conducted between 10^th^ to 30^th^ April 2020, during the lockdown stage [13]. Since during this special time it was impossible to conduct a community-based sampling survey, we decided to collect the data online. In this study, the community pharmacists working in Pakistan’s two provinces (Punjab and Khyber Pakhtunkhwa) were selected. A total registered pharmacist in Pakistan are 34000, about 29,000 of these are registered in Punjab (26,000) and Khyber Pakhtunkhwa (3,000) [14]. A study concluded that only 10% (n=2900) of registered pharmacists are working at community pharmacies in Pakistan [11].

### 2.2. Sample size

The minimum obligatory sample size calculated was 340 by using Raosoft sample size calculator [15], with a population size of 2900. Response distribution was assumed as 50%, power was kept at 80%, the margin of error 5%, and 95% confidence level was chosen for sample estimation.

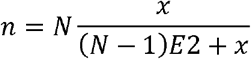

Where x is confidence interval, N is the population size, and E is the margin of error. Taking into consideration an additional 20% (n=68), for nonresponse, inappropriate responses, and error in questionnaire filling, a final sample size of 408 pharmacists will be required.

### 2.3. Data collection

A survey was started on 11 April 2020 and response acceptance was closed on 30 April 2020 when the required sample size was achieved. A two-page online questionnaire link was posted/reposted to various community pharmacists individually or in their groups via e-mails, WhatsApp, Instagram, and Facebook accounts. This questionnaire included a brief introduction to the context, purpose, procedures, anonymity, and confidentiality statements, as well as notes for completing the questionnaire. Participants had to answer a yes-no question to approve their willingness to participate deliberately. After the approval of the question, the participants were guided to complete the self-reporting questionnaire. Pharmacists working in hospitals, pharmaceutical, and marketing industries were excluded from this study.

### 2.4. Study questionnaire

A questionnaire was developed by the authors according to the COVID-19 guidelines of the international pharmaceutical federation (FIP) for pharmacists and the pharmacy workforce [8] and the national action plan for Coronavirus disease (COVID-19) Pakistan [16]. A pilot study was conducted on 40 pharmacists, 20 from each province. The reliability coefficient was estimated by using SPSS version 21 and Cronbach alpha found to be 0.745.

There were three sections of the questionnaire: demographics, basics questions and knowledge, attitudes, and practices (KAP). Demographic variables included age, gender, work areas (province name), marital status, pharmacy ownership, community pharmacist experience, and qualifications. Basics questions contained 3 items related to the source of knowledge, need for more education and training, and the level of concern about COVID-19.

The third part related to KAP was divided into three parts. The knowledge related portion had 24 questions about COVID-19 and were answered on a yes/no basis option. 1 point was assigned to the correct option answer and 0 point was assigned to the wrong answer. The total score ranges from 0-24 and a score of ≤19 indicted poor knowledge while a score of ≥20 (more than 80% of total score) demonstrated good knowledge.

The attitude/perception section consisted of 13-items. Responses were recorded on a five-point “Likert scale” as strongly disagree=1, disagree=2, uncertain=3, agree=4, strongly agree=5. Total score ranges from 1-65; a score of ≤ 52 indicated poor attitude and a score of ≥53 (more than 80% of total score) set for a good attitude.

The last part of the questionnaire comprised of 8-questions related to the practice of community pharmacists and each question scored as ‘yes’ (1-point), ‘no’ (0-point), and ‘sometimes’ (0-point). A total score ranges from 0-8 and a score of ≤ 6 demonstrated poor practice while a score of 7-8 (more than 75% of total score) indicated good practice.

### 2.5. Ethics

The study was carried out according to the declaration of Helsinki. This study was conducted in a strict locked down period and educational institutes were also closed; hence, the study protocol was approved by the ethical committee of the local teaching hospital (Reference number: 817/THQ/HR). The online questionnaire had a pre-face that describes the nature and purpose of the study and a consent part that ensures anonymity and voluntary participation of participants.

### 2.6. Statistical analysis

Statistical analysis was performed using SPSS version 21. Descriptive statistics were measured as frequencies and percentages for categorical variables. Chi-square tests used to check a difference in knowledge, attitude, and practice by participant’s characteristics. Multivariable binary logistic regression models were employed to find potential factors linked with good knowledge, attitude, and practice. Results were stated as odds ratios accompanied by 95%-confidence interval. Pearson correlation tests were applied to determine the nature of correlation among knowledge, attitude, and practice sections. A p-value of less than 0.05 was considered as a level of significance.

## 3. Results

A total of 393 participants (15 responses were excluded due to error in filling questionnaire/missed information) were included in the final analysis, of which half (52.2%, n=205) of respondents were male, 71% (n=279) were single and similar proportion (72.3%, n=284) were working as an employee in pharmacy. The majority of pharmacists (67.4%, n=265) had an experience of five years or less, 69% of participants had a pharm-d degree and 85.8% (n=337) wished to obtain more education and training regarding COVID-19 (Table 1).

**Table 1.** Sample Characteristics.

| Variable | Frequencies | Percentages |
| --- | --- | --- |
| **Gender** |  |  |
| Male | 205 | 52.2 |
| Female | 188 | 47.8 |
| **Age** |  |  |
| ≤25 years | 124 | 31.5 |
| ≥26 years | 269 | 68.5 |
| **Marital Status** |  |  |
| Married | 114 | 29.0 |
| Single | 279 | 71.0 |
| **Pharmacy ownership** |  |  |
| Owned | 67 | 17.0 |
| Partnership | 42 | 10.7 |
| Employed | 284 | 72.3 |
| **Experience** |  |  |
| ≤5 years | 265 | 67.4 |
| >5 Years | 128 | 32.6 |
| **Education?** |  |  |
| B-Pharm | 6 | 1.5 |
| Pharm-D | 271 | 69.0 |
| Master/M.Phil. | 107 | 27.2 |
| PhD | 9 | 2.3 |
| **Need of education?** |  |  |
| Yes | 337 | 85.8 |
| No | 56 | 14.2 |
| **Level of concern?** |  |  |
| Very High Concern | 231 | 58.8 |
| High Concern | 122 | 31.0 |
| Moderate Concern | 35 | 8.9 |
| Low concern | 3 | 0.8 |
| Very Low Concern | 1 | 0.3 |

The majority of the respondents utilized social media (45.29%, n=178) as the main source to seek information regarding COVID-19 followed by research articles (18.57%, n=73) and the ministry of health website (14.5%, n=57). The various information sources utilized by community pharmacists are summarized in figure 1.

**Figure 1:**
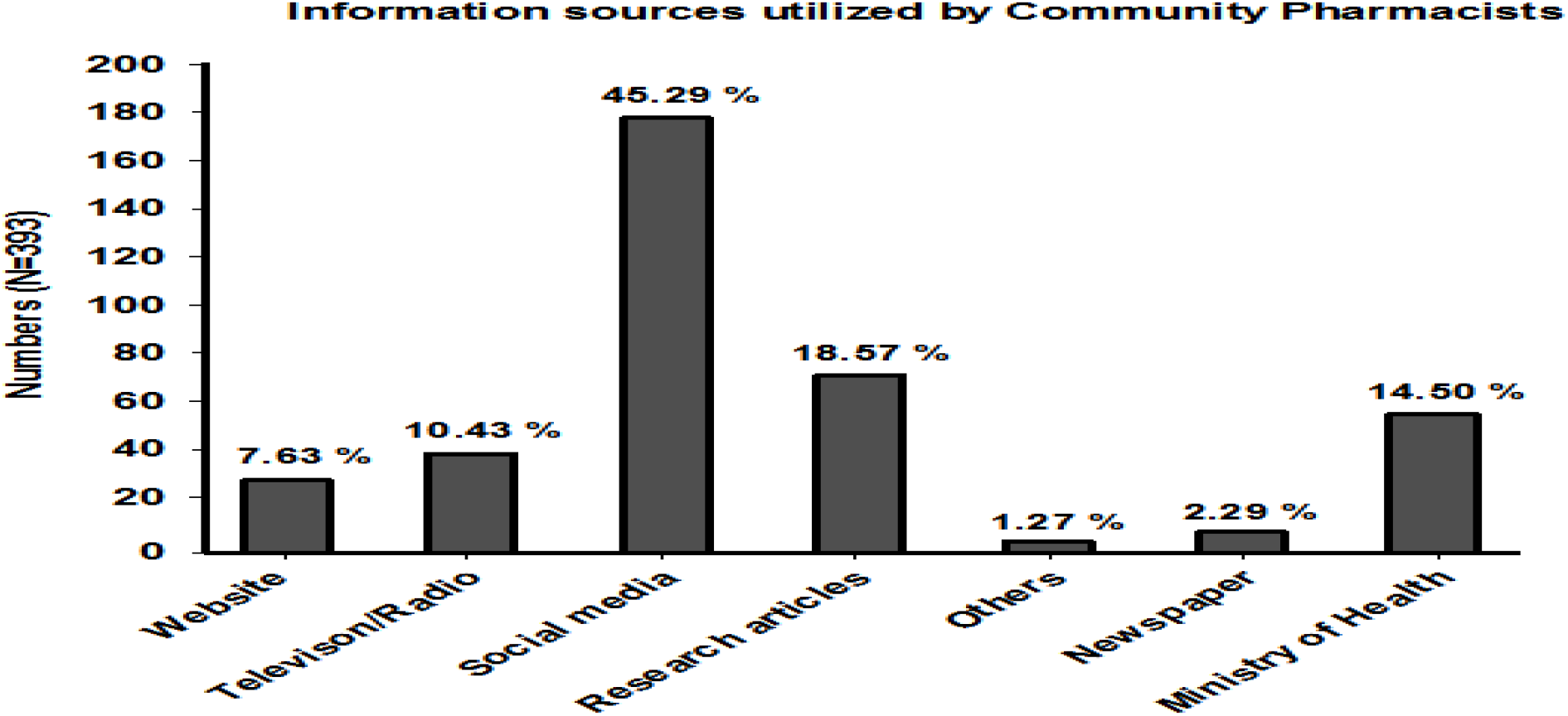
The various information sources utilized by community pharmacists (n=393)

### 3.1. Knowledge among Community pharmacists regarding COVID-19

Mixed responses were obtained regarding 24 knowledge items. Overall, the majority of respondents (71.5%, n=281) had good knowledge regarding COVID-19. More than 90% of participants had good knowledge regarding nature, symptoms, risky groups, and transmission of COVID-19. Additionally, 81.6% of participants were aware that there is no vaccine available in the market and 98.20% reported that the crowd is one of the major factors in the spread of disease. On the other hand, 30.80% didn’t knew that this is a zoonotic illness, and 28.8% reported that antibiotics are the first-line treatment. The responses obtained for knowledge items of the questionnaire are represents in supplementary file 1.

### 3.2. Attitude among Community pharmacists regarding COVID-19

All the participants responded to all 13 items on their attitude regarding COVID-19. Findings demonstrated that only 44% (n=173) of community pharmacists had a positive attitude towards COVID-19. Of the total participants, 65.40% strongly agreed that COVID-19 is a world health concern, less than half (47.60%) agreed that it is also a problem in Pakistan, 50.60% strongly agreed that pharmacists should avail themselves with updated knowledge, and more than 92% of the respondents were strongly agreed/agreed that community pharmacists could play an important role in this pandemic. On the other hand, 36.10% of the community pharmacists disagreed that the Pakistani populations have sufficient information, and 20.40% disagreed that government institutions can control the pandemic. The responses obtained for attitude items are presented in supplementary file 2.

### 3.3. Practice among Community pharmacists regarding COVID-19

More than half (57.3%, n=225) of pharmacists had followed good practices regarding COVID-19. The majority of respondents had good practice regarding each item with the highest practice showed in putting used tissue in the basket (91.90%), wearing a face mask (91.60%), washing hands (90.60%), and covering of eyes and nose with tissue (90.30%). A lower percentage of good practice was observed among community pharmacists in wearing protective gowns (57.50%) and in avoiding touching of eyes, nose, or mouth (74.60%). The responses obtained for practice assessing items of the questionnaire are listed in figure 2.

**Figure 2:**
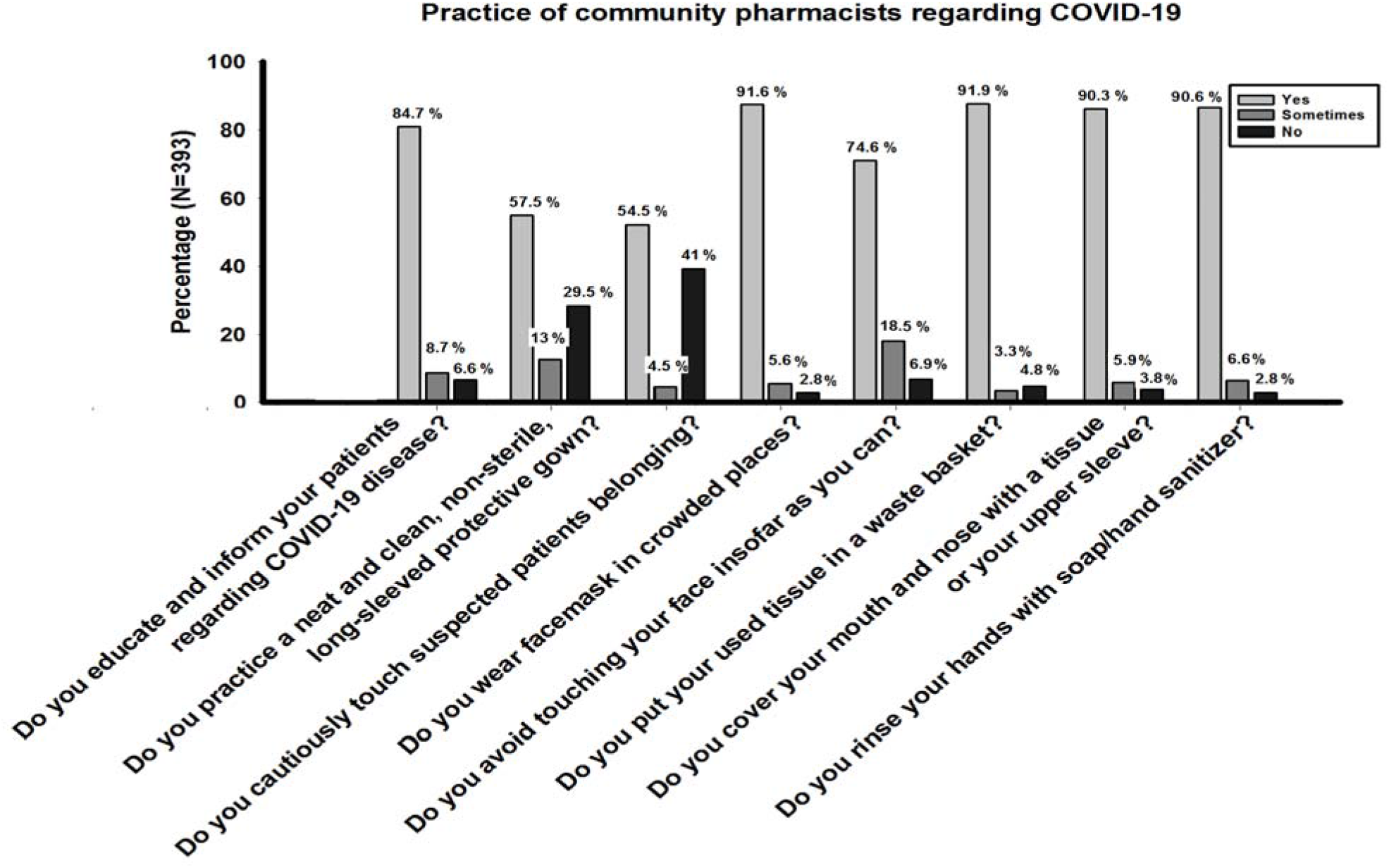
Practice among Community pharmacists regarding COVID-19.

### 3.4. The difference in knowledge, attitude, and practice among Community pharmacists about COVID-19

Chi-square tests were used to assess a difference in knowledge, attitude, and practice by sample characteristics. Findings revealed that community pharmacist’s knowledge regarding COVID-19 was significantly differed by gender (χ^2^ = 18.78, P<0.001), and level of concern (χ^2^ = 11.603, P=0.021) regarding COVID-19. As respondents who had very high concern have good knowledge compared to pharmacists who had less concern regarding the COVID-19 pandemic in Pakistan.

Results show that more than half (56%, n=220) of community pharmacists had a poor attitude regarding COVID-19. Factors that differentiate attitude status were found to be gender (χ^2^ = 4.789, P=0.033), education level (χ^2^ = 6.093, P=0.047) and level of concern (χ^2^ = 12.886, P=0.005).

Of 393 community pharmacists’, only 57.3% had good practice in following precautionary measures regarding COVID-19. Findings demonstrated that female pharmacists had good practice in following precautions compared to male counterparts (χ^2^ = 5.383, P=0.020). Other factors that significantly associated with practice status were age (χ^2^ = 5.322, P=0.025), and education level (χ^2^ = 9.831, P=0.020). The complete details are listed in table 2.

**Table 2.** Difference in Knowledge, attitude and practice by sample characteristics.

| Variables | Knowledge |  |  | Attitude |  |  | Practice |  |  |
| --- | --- | --- | --- | --- | --- | --- | --- | --- | --- |
| | Poor | Good | $\chi^2$ (P) | Poor | Good | $\chi^2$ (P) | Poor | Good | $\chi^2$ (P) |
|  | [n (%)] | [n (%)] |  | [n (%)] | [n (%)] |  | [n (%)] | [n (%)] |  |
| **Total** | 112 (28.5) | 281 (71.5) |  | 220 (56.0) | 173 (44.0) |  | 168 (42.7) | 225 (57.3) |  |
| **Gender** |  |  | 18.78 (<0.001) |  |  | 4.789 (0.033) |  |  | 5.383 (0.020) |
| Male | 39 (19.0) | 166 (81.0) |  | 104 (50.7) | 101 (49.3) |  | 99 (48.3) | 106 (51.7) |  |
| Female | 73 (38.8) | 115 (61.2) |  | 116 (61.7) | 72 (38.3) |  | 69 (36.7) | 119 (63.3) |  |
| **Age** |  |  | 0.775 (0.379) |  |  | 10.171 (0.001) |  |  | 0.049 (0.825) |
| ≤25 years | 39 (31.5) | 85 (68.5) |  | 84 (67.7) | 40 (32.3) |  | 52 (41.9) | 72 (58.1) |  |
| ≥26 years | 73 (27.1) | 196 (72.9) |  | 136 (50.6) | 133 (49.4) |  | 116 (43.1) | 153 (56.9) |  |
| **Marital Status** |  |  | 1.827 (0.172) |  |  | 0.166 (0.684) |  |  | 5.322 (0.025) |
| Married | 27 (23.7) | 87 (76.3) |  | 62 (54.4) | 52 (45.6) |  | 59 (51.8) | 55 (48.2) |  |
| Single | 85 (30.5) | 194 (69.5) |  | 158 (56.6) | 121 (43.4) |  | 109 (39.1) | 170 (60.9) |  |
| **Pharmacy ownership** |  |  | 0.894 (0.674) |  |  | 0.832 (0.660) |  |  | 0.700 (0.705) |
| Owned | 16 (23.9) | 51 (76.1) |  | 35 (52.2) | 32 (47.8) |  | 26 (38.8) | 41 (61.2) |  |
| Partnership | 12 (28.6) | 30 (71.4) |  | 22 (52.4) | 20 (47.6) |  | 17 (40.5) | 25 (59.5) |  |
| Employed | 84 (29.6) | 200 (70.4) |  | 163 (57.4) | 121 (42.6) |  | 125 (44.0) | 159 (56.0) |  |
| **Experience** |  |  | 0.013 (0.909) |  |  | 0.259 (0.611) |  |  | 2.136 (0.144) |
| ≤5 years | 76 (28.7) | 189 (71.3) |  | 146 (55.1) | 119 (44.9) |  | 120 (45.3) | 145 (54.7) |  |
| >5 Years | 36 (28.1) | 92 (71.9) |  | 74 (57.8) | 54 (42.2) |  | 48 (37.5) | 80 (62.5) |  |
| **Education level** |  |  | 1.760 (0.624) |  |  | 6.093 (0.047) |  |  | 9.831 (0.020) |
| B-Pharm | 3 (50.0) | 3 (50.0) |  | 5 (83.3) | 1 (16.7) |  | 3 (50.0) | 3 (50.0) |  |
| Pharm-D | 78 (28.8) | 193 (71.2) |  | 154 (56.8) | 117 (43.2) |  | 107 (39.5) | 164 (60.5) |  |
| Master/M.Phil. | 28 (26.2) | 79 (73.8) |  | 59 (55.1) | 48 (44.9) |  | 57 (53.3) | 50 (46.7) |  |
| PhD | 3 (33.3) | 6 (66.7) |  | 2 (22.2) | 7 (77.8) |  | 1 (11.1) | 8 (88.9) |  |
| **Need of education?** |  |  | .392 (0.531) |  |  | 1.828 (0.176) |  |  | 0.320 (0.572) |
| Yes | 98 (29.1) | 239 (70.9) |  | 184 (54.6) | 153 (45.4) |  | 146 (43.3) | 191 (56.7) |  |
| No | 14 (25.0) | 42 (75.0) |  | 36 (64.3) | 20 (35.7) |  | 22 (39.3) | 34 (60.7) |  |
| **Level of concern?** |  |  | 11.603 (0.021) |  |  | 12.886 (0.005) |  |  | 4.576 (0.334) |
| Very High Concern | 68 (29.4) | 163 (70.6) |  | 113 (48.9) | 118 (51.1) |  | 97 (42.0) | 134 (58.0) |  |
| High Concern | 29 (23.8) | 93 (76.2) |  | 80 (65.6) | 42 (34.4) |  | 48 (39.3) | 74 (60.7) |  |
| Moderate Concern | 11 (31.4) | 24 (68.6) |  | 24 (68.6) | 11 (31.4) |  | 19 (54.3) | 16 (45.7) |  |
| Low concern | 3 (100.0) | 0 (0.0) |  | 1 (33.3) | 2 (66.7) |  | 2 (66.7) | 1 (33.3) |  |
| Very Low Concern | 1(100.0) | (0.0) |  | 1 (100.0) | (0.0) |  | 1 (100.0) | 0 (0.0) |  |

### 3.5. Factors associated with good knowledge, attitude, and practice about COVID-19

Binary logistic regression analysis was performed to evaluate the substantial determinants of good knowledge, attitude, and practice about COVID-19 amid community pharmacists in Pakistan. Findings indicated that females had 0.243 times lower odds (OR: 0.243, 95% CI: 0.135-0.436, P<0.001) of good knowledge as compared to male counterparts. Results revealed that age of ≥26 years (OR:2.046, 95% CI:1.214-3.450. P=0.007), Ph.D. degree level (OR: 14.16, 95%CI:0.875-22.95, P=0.042), and good knowledge (OR: 2.045, 95%CI: 1.233-3.391, P=0.006) were the significant determinants of a good attitude. Similarly, community pharmacist who had experience of >5 years (OR: 1.505, 95%CI: 0.944-2.40, P=0.048), PhD degree (OR: 6.574, 95%CI: 0.362-11.942, P=0.021), good knowledge (OR: 2.448, 95%CI: 1.485-2.376, P<0.001) and good attitude (OR: 1.518, 95%CI: 0.970-2.376, P=0.048) had higher odds of good practice compared to reference categories (Table 3).

**Table 3.** Regression analysis for factors associated with Good knowledge, Attitude and Practice regarding COVID-19.

| Variables | Knowledge |  | Attitude |  | Practice |  |
| --- | --- | --- | --- | --- | --- | --- |
|  | OR (95% CI) | P | OR (95% CI) | P | OR (95% CI) | P |
| **Gender** |  |  |  |  |  |  |
| Male | 1.00 | - | 1.00 | - | 1.00 | - |
| Female | 0.243 (0.135- 0.436) | **<0.001** | 0.859 (0.507-1.455) | 0.572 | 1.79 (1.038-3.086) |  |
| **Age** |  |  |  |  |  |  |
| ≤25 years | Reference | - | 1.00 | - | 1.00 | - |
| ≥26 years | 1.783 (1.425-2.282) | 0.281 | 2.046 (1.214-3.450) | **0.007** | 1.395 (0.822-2.368) | 0.217 |
| **Marital Status** |  |  |  |  |  |  |
| Married | 1.00 | - | 1.00 | - | 1.00 | - |
| Single | 1.022 (0.562-1.857) | 0.944 | 1.181 (0.708-1.969) | 0.525 | 1.607 (0.955-2.704) | 0.074 |
| **Pharmacy ownership** |  |  |  |  |  |  |
| Owned | 1.00 | - | 1.00 | - | 1.00 | - |
| Partnership | 0.785 (0.180-1.664) | 0.991 | 1.039 (0.440-2.453) | 0.931 | 0.902 (0.379-2.148) | 0.816 |
| Employed | 0.866 (0.426-1.760) | 0.690 | 1.006 (0.550-1.841) | 0.984 | 0.694 (0.377-1.277) | 0.240 |
| **Experience** |  |  |  |  |  |  |
| ≤5 years | 1.00 | - | 1.00 | - | 1.00 | - |
| >5 Years | 1.107 (0.671-1.826) | 0.690 | 1.007 (0.638-1.590) | 0.976 | 1.505 (0.944-2.40) | **0.048** |
| **Education?** |  |  |  |  |  |  |
| B-Pharm | 1.00 | - | 1.00 | - | 1.00 | - |
| Pharm-D | 2.307 (0.207-25.735) | 0.390 | 4.268 (0.440-41.41) | 0.211 | 1.928 (0.163-5.288) | 0.133 |
| Master/M.Phil. | 2.707 (0.465-1.748) | 0.281 | 4.663 (0.374-35.83) | 0.265 | 2.618 (1.106-4.621) | 0.193 |
| PhD | 3.541 (0.623-20.139) | 0.497 | 14.16 (0.875-22.95) | **0.042** | 6.574 (0.362-11.942) | **0.021** |
| **Need of education?** |  |  |  |  |  |  |
| Yes | 1.00 | - | 1.00 | - | 1.00 | - |
| No | 1.933 (0.942-3.970) | 0.073 | 0.626 (0.333-1.176) | 0.145 | 1.107 (0.585-2.093) | 0.754 |
| **Level of Concern?** |  |  |  |  |  |  |
| Very High Concern | 1.00 | - | 1.00 | - | 1.00 | - |
| High Concern | 0.991 (0.496-1.793) | 0.220 | 3.496 (0.308-3.800) | **0.004** |  | 0.491 |
| Moderate Concern | 0.878 (0.496-1.793) | 0.278 | 3.443 (0.200-3.980) | **0.044** |  | 0.528 |
| Low concern | 0.799 (0.013-1.071) | 0.999 | 2.498 (0.198-3.193) | 0.479 |  | 0.453 |
| Very Low Concern | 0.782 (0.019-1.028) | 0.999 | 2.368 (0.143-3.872) | 0.899 |  | 0.899 |
| **Knowledge** | - | - |  |  |  |  |
| Poor | - | - | 1.00 | - | 1.00 | - |
| Good | - | - | 2.045 (1.233-3.391) | **0.006** | 2.448 (1.485-2.376) | **<0.001** |
| **Attitude** | - | - | - | - |  |  |
| Poor | - | - | - | - | 1.00 | - |
| Good | - | - | - | - | 1.518 (0.970-2.376) | **0.048** |

Pearson correlation tests showed a linear statistically significant positive correlation amid knowledge, attitude and practice scores as follows: knowledge-attitude (r = 0.274, P<0.001), attitude-practice (r = 0.170, P=0.001), and knowledge-practice (r = 0.341, P<0.001) (Table 4).

**Table. 4.** Correlation between scores of knowledge, attitude, and practice.

| **Variable** | **Correlation Coefficient** | **P-Value** |
| --- | --- | --- |
| Knowledge-Attitude | 0.274* | **<0.001** |
| Attitude-Practice | 0.170* | 0.001 |
| Knowledge-Practice | 0.341* | <0.001 |

\* Correlation significant at 0.05 level (2 tailed).

## 4. Discussion

Pharmacists play a pivotal role in the provision of drug-related information to patients, caregivers, and healthcare practitioners (HCPs) [17]. The FIP emphasizes the active role of pharmacists in the community and hospital setups in preventing the spread of COVID-19 [8]. Community pharmacists are easily available and readily accessible to the general population. They are the first point of contact for all those individuals who need information or guidance about medicines [1, 2, 6, 8]. According to the best of our knowledge, this is the first-ever research study that thoroughly assessed the knowledge, attitudes, and practice of community pharmacists towards COVID-19 in Pakistan.

In this study, it is revealed that the majority of the respondents used social media (45.3%) followed by research articles (18.57%) as their primary source to get information about COVID-19. These results were in agreement with findings of Basheti et al. (59.5%) [2], Saqlain et al (87.68%) [18], Giao et al (91.1%) [19] and Bhagavathula et al (61.0%) [20]. Whereas, Kara et al. reported that the participants generally used television, newspapers, and the internet as a principal source to get information about COVID-19 [17]. Moreover, our study showed that research articles were also searched by community pharmacists as a source of information because valid and satisfactory information can be obtained from standard research articles. On the other side, community pharmacists can be misguided by the false information that is available on the internet and circulates through social media. Therefore, community pharmacists and other HCPs should carefully evaluate sources of COVID-19 information and use only standard and authentic content to seek information [17, 20].

A good knowledge, positive attitude, and practices among community pharmacists regarding basic precautionary measures (e.g. wearing protective clothing, goggles, face mask, and gloves) are important to deal with general community as it decreases the chances of transmission. Moreover, the current pandemic nature of the COVID-19 has made it mandatory for a community pharmacist to increase their precautions according to the critical situation. Also, they must make attempts in implementing recommendations and following relevant hygienic conditions. On the other hand, good knowledge and practice of community pharmacists in complying precautionary measures not only create awareness among patients but also gives an important message in society [2, 8, 17].

The majority of respondents (71.5%) in this study had good knowledge regarding COVID-19. Other findings revealed that the knowledge of community pharmacists was significantly different by the level of concern and gender. The recent studies conducted in Turkey (90%) among pharmacist [17], in Iran (56.5%) among nurses [20] and in Pakistan (93.2%) [18] and China (88.4%) among HCPs [19], showed different knowledgeable rate about COVID-19. The results of this study provide confidence in terms of the community pharmacist knowledge regarding the signs, transmission, and preventive measures of COVID-19. This is of greater concern in the present situation because there is no vaccine available and work continues, so that health practitioners, including community pharmacists and paramedics, must be familiar with all the latest developments and should take precautionary steps in the prevention and treatment of diseases [2, 6, 18].

On the other hand, 30.80% didn’t knew this is a zoonotic illness in this study. Similar findings were reported by Kara et al. (30%) [17] and Bhagavathula et al (20%) [20]. However, it is reported that COVID-19 was closely linked to a wet market in China, and other viral diseases such as SARS, MERS, and Ebola emerged from zoonotic origins [1, 2, 21]. Furthermore, about 28.8% reported incorrectly that antibiotics are the first-line treatment. At present, the treatments of patients with COVID-19 are mainly repurposing the available therapeutic drugs and based on symptomatic conditions [22]. Antibiotics are not a first-line effective choice in preventing or treating the COVID-19 and misusing antibiotics can result in the development of antimicrobial resistance [23, 24].

Regression analysis revealed that females had lower odds (OR: 0.243, P<0.001) of good knowledge compared to male counterparts (OR: 1.00). However, Zhong et al [12] showed that male gender (vs. female, β: −0.284, P<0.001) were significantly associated with lower knowledge score. Significantly higher knowledge scores among males than females may be related to a higher level of education among males in Pakistan [25]. The finding suggests that intervention via health education can be more efficient if it targets specific demographic groups, such as, the COVID-19 knowledge may be markedly increased if the health education programs are certainly designed for females and less educated peoples [12, 18].

The majority of the community pharmacists (92.4%) perceived that they could play an important role in this pandemic. Similar findings were also reported by Basheti et al. in Jorden (70%) [2]. Nearly 36.6% of the participants perceived that government health care institutes in Pakistan can control COVID-19, while 28.8% disagreed. A recent study conducted in China reported that the majority of the respondents (97.1%) were confident and agreed that their government can win the fight against COVID-19 [12]. Lack of confidence among the participants in our study may be due to the lack of basic health facilities, recommended policies, unavailability of proper medical equipment, political up-downs, financial crisis, and de-stabilized economic condition of Pakistan to cope with such outbreak [10]. Additionally, about 36.10% of the respondents disagreed that the Pakistani populations have sufficient information. This may be due to the perception that the Pakistani population has limited access to the internet, limited health information resources and mostly living in rural areas.

This study demonstrated that only 44% of the participants had a positive attitude towards COVID-19. The attitude status of community pharmacists about COVI-19 needs further improvement. These results can be used by the general public, policy-makers, and health workers to observe the targeted population for the prevention of COVID-19 and health education. Regression analysis revealed that pharmacists age ≥26 years had higher odds (OR:2.406, P<0.001) of good attitude which is in line with a Turkish study which also reported that age was a factor that influenced the participants’ attitudes towards COVID-19 infection [17]. On contrary, a study conducted in China did not reveal any association of age, gender, and experience with attitude level [19]. This variation could be possibly explained by the geographical difference between these countries.

Findings also showed that pharmacists with a higher-level degree (doctor of philosophy; Ph.D.) had better (OR:14.16, P=0.048) attitude compared to other educational levels. These findings are also supported by Naser et al. [26] and Zhong et al. [12]. A higher educational degree appreciably increases knowledge and ultimately a positive attitude. Targeted educational programs are needed to raise awareness about COVID-19. Such programs need to be tailored to young individuals and particularly those with lower education levels [26, 27].

A pharmacist who had a higher level of concern demonstrated a good attitude compared to less concerned pharmacists. Of note that, good knowledge found to be a potential predictor of positive attitude (OR: 2.045, P=0.006). This was also found in correlation as knowledge is positively correlated with attitude (r=0.274, P<0.001). Our finding was supported by the studies conducted in China [12], Iran [20], and Thailand [28]. Therefore, continuous educational programs and periodic training may be an effective intervention to improve the knowledge, level of concern, and ultimately attitude of the community pharmacist about COVID-19 pandemic.

More than half (57.3%) of the respondents showed good practice towards COVID-19. The overall highest practice observed amid participants were regarding throwing the used tissue in the trash (91.90%) followed by wearing a face mask (91.6%), and washing hands (90.6%). A recently conducted study showed that only 8.9% of the pharmacist’s used face masks and 84.8% washed their hands at the workplace [17]. The spread of COVID-19 can be prevented if the health care workers wash their hands with soap and water at specific times and maintain an excellent hygiene condition. However, the least practice observed among participants were wearing protective gowns (57.5%). This result is of special concern because good knowledge with poor practice, not only increases the transmission of infection but also increases the morbidity and mortality rates in the community [29]. It was noted that personal protective equipment (PPE) decreases the transmission of microbes in the hospital setup and also protects peoples from infections in the community [30]. Therefore, it is crucial between various community pharmacists and other HCPs to follow the practice guidelines recommended by the national institute of health (NIH), Islamabad, Pakistan, centers for disease control and prevention (CDC), and world health organization WHO regarding the COVID-19 infection. It further suggests the community pharmacists and other healthcare workers to boost their knowledge and attitude that will ultimately translate into good practice.

Findings showed that community pharmacists with good knowledge (OR:2.44, P<0.01) and positive attitude (OR: 1.15, P=0.048) demonstrated good practices in following precautionary measures. The studies conducted by Kara et al. [17], Saqlain et al. [18], and Naser et al. [26] indicated that pharmacists with good knowledge had a good attitude and showed good practices. Therefore, adequate knowledge is crucial and could be improved via an extensive educational program for better understanding and improved practices [31].

### Strengths and Limitations

This study was carried out in less explored areas where scare literature was present about the current topic. The study respondents were community pharmacists, which are educated and professionals enough to answer with honesty and responsibility. This study presents the current status of the community pharmacist knowledge, which is the first basic and important aspect of a successful response to an epidemic. This study was a computer-generated survey and is free of errors as compared to hand filled proforma. Moreover, the study describes the community pharmacist knowledge, attitude, and practices towards COVID-19 in detail, which suggests that the health ministry and related departments should focus independently. WHO published materials that were used in the development of the questionnaire and a two-step data validation technique was used, hence all these factors increase the reliability of the questionnaire.

This study has a few limitations. It is a cross-sectional survey and carried out during the locked down period when educational institutes were closed, hence approval from the institutional review board was not approached. Additionally, it is an online survey, where responses primarily depend upon honesty and are partly affected by recall ability therefore chances of biasness may be there. Finally, this study is the unstandardized and inadequate assessment of attitudes and practices towards COVID, which should be established through focus group discussion and comprehensive interviews and constructed as multi-dimensional measures.

### Conclusion

The community pharmacists had good knowledge but had a poor attitude and practice towards the COVID-19. The majority of the community pharmacists perceived that they can play an important role in this pandemic. This study also highlighted the disparity in some aspects of knowledge, attitude, and practice that must be addressed in future educational, awareness, and counselling programs. It is important for all the HCPs including pharmacists to have standard authentic information about the COVID-19 and to further convey this knowledge and belief to the community. Future studies are required to evaluate the knowledge, attitudes, and practices of other HCPs and other segments of society. This study recommends the health ministry and other associated authorities to promote awareness about COVID-19 and its related symptoms with a comprehensive training program. These programs should be consisting of better-structured targeting not only for medical doctors but also pharmacists, nurses, and other paramedical staff to build equilibrium in clinical knowledge about COVID-19.

## Data Availability

All data available inside the manuscript

## Acknowledgements

The authors would like to thank to all the participants for their help.

## Authors’ contributions

KM, MS and ZK involve in conceptualization, project administration, resources, investigation, methodology, data curation, writing – original draft, Writing – review & editing. AH, NA, MFU done data curation, software, writing – review & editing. SK, MK, HR, SK involved in critical help in writing, manuscript writing process and layout designing. MIK, FK and YH provided critical input regarding data analysis at every step of the manuscript writing process and input in formulating the manuscript draft. All authors approved and agreed to the submission of manuscript.

## Supplementary Files

**Supplementary file 1:**
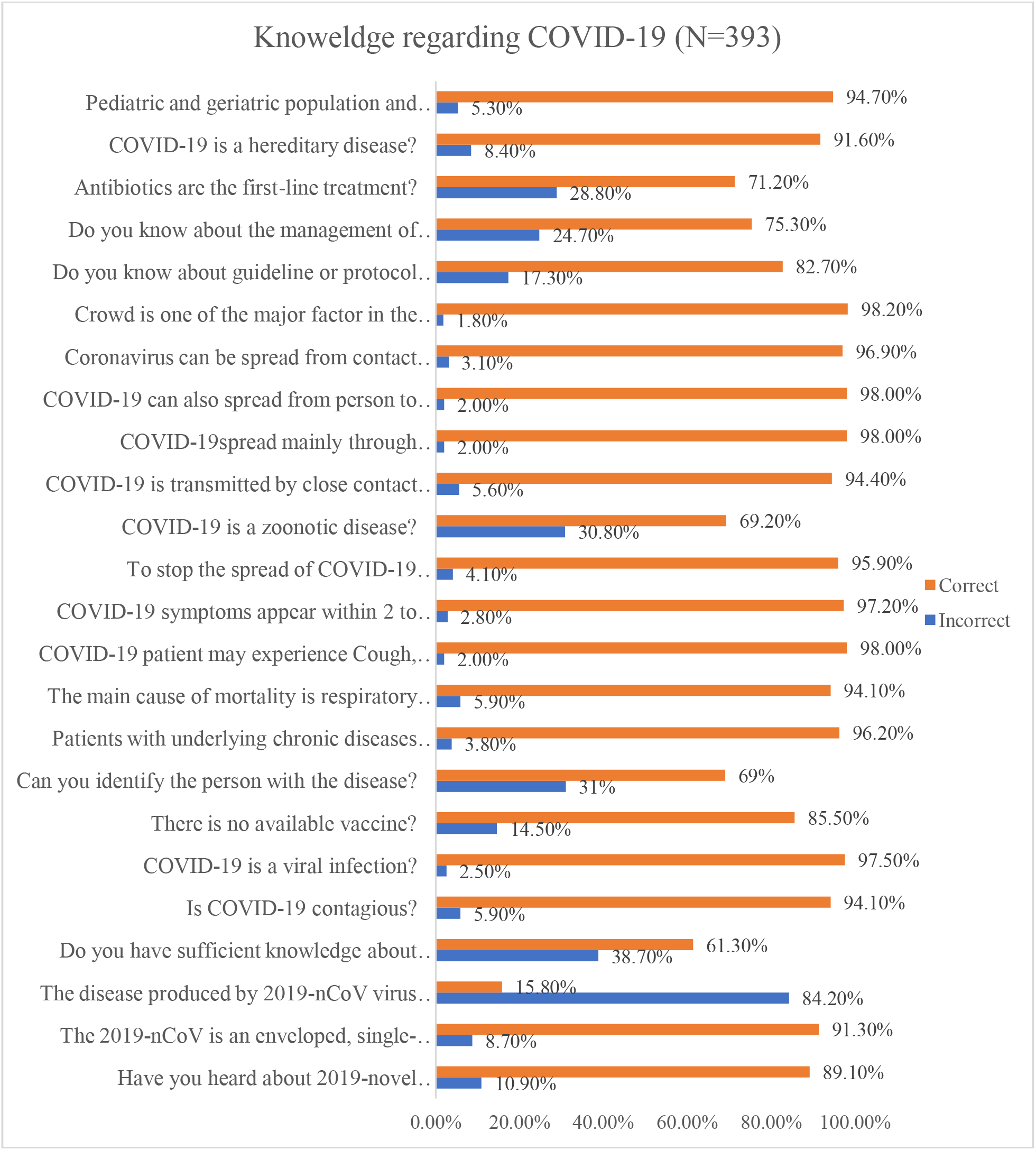
Knowledge among Community pharmacists regarding COVID-19.

**Supplementary file 2:**
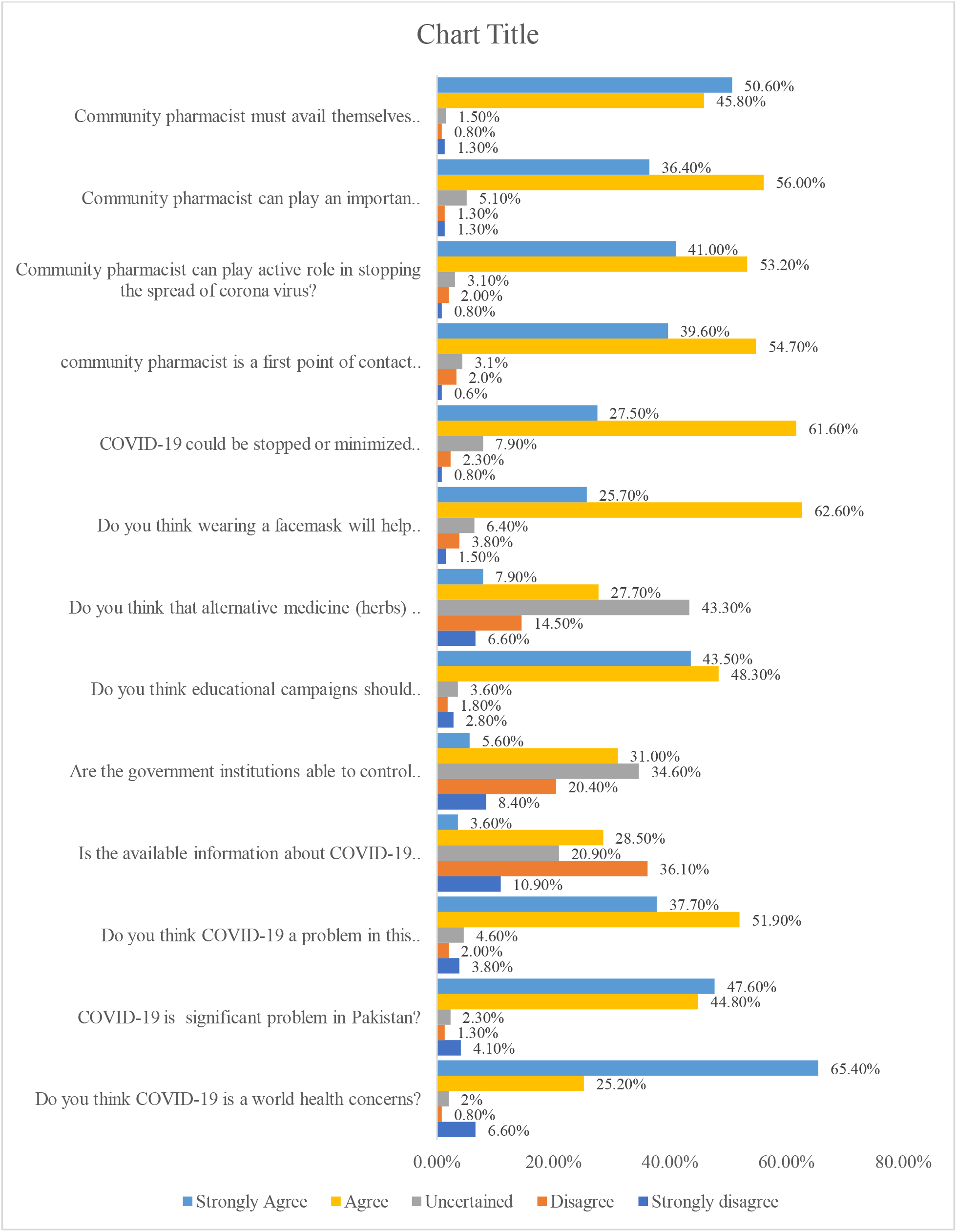
The attitude among Community pharmacists regarding COVID-19.

## References

1. Khan Z, Muhammad K., Ahmed A. Rahman H. Coronavirus outbreaks: prevention and management recommendations. Drugs Ther Perspect 36, 215–217 (2020). https://doi.org/10.1007/s40267-020-00717-x.

2. Basheti IA, Nassar R, Barakat M, Alqudah R, Abufarha R, Mukattash TL, et al. Pharmacists’ readiness to deal with the coronavirus pandemic: Assessing awareness and perception of roles. Res Social Adm Pharm. 2020. In Press. https://doi.org/10.1016/j.sapharm.2020.04.020.

3. Paules CI, Marston HD, Fauci AS. Coronavirus Infections-More Than Just the Common Cold. JAMA. 2020;323(8):707–708. DOI:10.1001/jama.2020.0757.

4. World Health Organization (WHO). Coronavirus disease 2019 (COVID-19) situation report—50. 2020. https://www.who.int/docs/default-source/coronaviruse/situation-reports/20200310-sitrep-50-covid-19.pdf?sfvrsn=55e904fb_2. Accessed 10 May 2020.

5. World Health Organization. Coronavirus disease 2019 (COVID-19) situation report: 119. Available from: https://www.who.int/docs/default-source/coronaviruse/situation-reports/20200518-covid-19-sitrep-119.pdf?sfvrsn=4bd9de254. Accessed 19 May 2020.

6. Dawoud D. Emerging from the other end: Key measures for a successful COVID-19 lockdown exit strategy and the potential contribution of pharmacists. Res Social Adm Pharm. 2020. In Press. https://doi.org/10.1016/j.sapharm.2020.05.011.

7. Ung COL. Community pharmacist in public health emergencies: Quick to action against the coronavirus 2019-nCoV outbreak. Res Social Adm Pharm. 2020; 16(4):583–586. https://doi.org/10.1016/j.sapharm.2020.02.003.

8. International Pharmaceutical Federation (FIP). Covid-19: guidelines for pharmacists and the pharmacy workforce. Updated 26 March 2020. Available from: https://www.fip.org/files/content/priority-areas/coronavirus/COVID-19-Guidelines-for-pharmacists-and-the-pharmacy-workforce.pdf.

9. Cattani M. Global coalition to accelerate COVID-19 clinical research in resource-limited settings. Lancet 2020. Doi: https://doi.org/10.1016/S0140-6736(20)30798-4. In Press.

10. Ul-Haq Z, Shah BH, Ardakani M, Khan SA, Muhammad S, Farooq S, et al. Health system preparedness in Pakistan for crisis management: a cross-sectional evaluation study. East Mediterr Health J. 2019;25(8):553–561. Doi: https://doi.org/10.26719/emhj.18.072.

11. Azhar S, Hassali MA, Taha A, Khan SA, Murtaza G, Hussain I. Evaluation of the perception of community pharmacists regarding their role in Pakistan’s healthcare system: a qualitative approach. Trop J Pharm Res. 2013;12(4):635–639.

12. Zhong BL, Luo W, Li HM, Zhang QQ, Liu XG, Li WT, Li Y. Knowledge, attitudes, and practices towards COVID-19 among Chinese residents during the rapid rise period of the COVID-19 outbreak: a quick online cross-sectional survey. International journal of biological sciences. 2020;16(10):1745.

13. Gardaworld news. “Pakistan: Government extends nationwide lockdown until April 30”. https://www.garda.com/crisis24/news-alerts/332256/pakistan-government-extends-nationwide-lockdown-until-april-30-update-18.

14. Pharmacy Council of Pakistan. 2020. Available from: https://www.pharmacycouncil.org.pk.

15. Raosoft sample size calculator. Available from: http://www.raosoft.com/samplesize.html.

16. National Action Plan for Corona virus disease (COVID-19) Pakistan. Updated 13 March 2020. Available from: https://www.nih.org.pk/wp-content/uploads/2020/03/CQVID-19-NAP-V2-13-March-2020.pdf.

17. Kara E, Demirkan K, Unal S. Knowledge and attitudes of hospital pharmacist about COVID-19) Turk J Pharm Sci 2020. DOI: 10.4274/tjps.galenos.2020.72325. [Ahead of Print].

18. Saqlain M, Munir MM, Rehman SU, Gulzar A, Naz S, Ahmed Z, Tahir AH, et al. Knowledge, attitude, practice and perceived barriers among healthcare professionals regarding COVID-19: A Cross-sectional survey from Pakistan. Journal of Hospital Infection. 2020. In Press. https://doi.org/10.1016/j.jhin.2020.05.007.

19. Giao H, Thi N, Han N, Khanh T Van, Ngan VK, Tam V Van, et al. Knowledge and attitude toward COVID-19 among healthcare workers at Knowledge and attitude toward COVID-19 among healthcare workers at District 2 Hospital, Ho Chi Minh City. Asian Pac J Trop Med 2020; 13:1–6. https://doi.org/10.4103/1995-7645.280396.

20. Bhagavathula AS, Aldhaleei WA, Rahmani J, Mahabadi MA, Bandari DK. Knowledge and Perceptions of COVID-19 Among Health Care Workers: Cross-Sectional Study. JMIR Public Health Surveill. 2020; 6(2):e19160. doi: 10.2196/19160. MedRxiv 2020:2020.03.09.20033381. https://doi.org/10.1101/2020.03.09.20033381.

21. Zhu N, Zhang D, Wang W, Li X, Yang B, Song J, et al. A Novel Coronavirus from Patients with Pneumonia in China, 2019. N Engl J Med. 2020 Feb 20;382(8):727–733. doi: 10.1056/nejmoa2001017.

22. Khan Z, Karata§ Y, Rahman, H. Anti COVID-19 Drugs: Need for More Clinical Evidence and Global Action. Adv Ther (2020). https://doi.org/10.1007/s12325-020-01351-9].

23. Wu R, Wang L, Kuo HD, Shannar A, Peter R, Chou PJ, et al. An Update on Current Therapeutic Drugs Treating COVID-19. Curr Pharmacol Rep (2020). https://doi.org/10.1007/s40495-020-00216-7

24. Xiao Y, Torok ME. Taking the right measures to control COVID-19. The Lancet Infectious Diseases. 2020;20(5): 523–524. https://doi.org/10.1016/S1473-3099(20)30152-3].

25. Pakistan Facts and Figures. 2018. Available from: https://centralasiainstitute.org/pakistan-facts-and-figures/.

26. Naser AY, Dahmash EZ, Alwafi H, Alsairafi ZK, Rajeh AMA, Alhartani YJ, et al. Knowledge and practices towards COVID-19 during its outbreak: a multinational cross-sectional study. medRxiv. 2020. https://doi.org/10.1101/2020.04.13.20063560.

27. Jamal A, Startsman K, Guy J, Pierce R, Ahmad A, Chang J, et al. Assessing Knowledge and Attitude about ebola in the US: a Cross Sectional Survey. Value Heal. 2015;18:A1–A307.

28. Srichan P, Apidechkul T, Tamornpark R, Yeemard F, Khunthason S, Kitchanapaiboon S, et al. Knowledge, Attitude and Preparedness to Respond to the 2019 Novel Coronavirus (COVID-19) Among the Bordered Population of Northern Thailand in the Early Period of the Outbreak: A Cross-Sectional Study. 2020. Available at SSRN: https://ssrn.com/abstract=3546046 or http://dx.doi.org/10.2139/ssrn.3546046.

29. Albarrak AI, Mohammed R, Al Elayan A, Al Fawaz F, Al Masry M, Al Shammari M, et al. Middle East Respiratory Syndrome (MERS): Comparing the knowledge, attitude and practices of different health care workers. J Infect Public Health 2019;617:6–13. https://doi.org/10.1016/j.jiph.2019.06.029.

30. Centers for Disease Control and Prevention (CDC). Personal protective equipment. 2018. Available from: https://www.cdc.gov/niosh/ppe/.

31. Athiyah U, Setiawan CD, Nugraheni G, Zairina E, Utami W, Hermansyah A. Assessment of pharmacists’ knowledge, attitude and practice in chain community pharmacies towards their current function and performance in Indonesia. Pharm Pract (Granada). 2019; 17(3): 1518. doi:10.18549/PharmPract.2019.3.1518.

